# Yellow fever virus resurgence in São Paulo State, Brazil, 2024-2025

**DOI:** 10.1101/2025.03.14.25323956

**Authors:** Mariana Sequetin Cunha, Juliana Mariotti Guerra, Márcio Junio Lima Siconelli, Benedito Antonio Lopes da Fonseca, Jessica Caroline de Almeida Dias, Gisele Dias de Freitas, Ester Sabino, Erika R. Manulli, Geovana M. Pereira, Ian N. Valença, Patrícia Sayuri Silvestre Matsumoto, Nuno R. Faria, Natália Coelho Couto de Azevedo Fernandes

**Author notes:** Correspondence to, Avenida Dr. Arnaldo, 355, Center of Virology, Adolfo Lutz Institute, São Paulo, Brazil.

## Abstract

Yellow fever virus (YFV) was detected in two distinct geographic locations in São Paulo State, Brazil, between September 2024 and February 2025. Phylogenetic analysis of six new genomes revealed a re-introduction in 2022 from Midwest Brazil followed by persistence in São Paulo state. Continued surveillance in neotropical primates is required to prevent cases in humans.

## Text

Yellow fever (YF) is a severe disease caused by *Orthoflavivirus flavi* (formerly known as Yellow fever virus, YFV), a member of the *Flaviviridae* family. It remains endemic in parts of Africa and the Americas, posing an ongoing public health threat (1). In Brazil, the urban cycle of YF, transmitted by *Aedes aegypti* mosquitoes, was eradicated in 1942. Since then, YFV has been maintained in a sylvatic cycle involving several neotropical primate species (NTPs) and forest canopy-dwelling mosquitoes, primarily *Haemagogus* spp. and *Sabethes* spp. (2) Consequently, YF surveillance relies on the confirmation of epizootic events through virus detection via RT-qPCR and/or immunohistochemistry, following the guidelines of the Brazilian Ministry of Health (3). Despite the availability of the live attenuated 17DD vaccine, from mid-2016 until late 2018, Brazil experienced one of the largest YF outbreaks in recent decades, predominantly affecting southeastern Brazil (4–7). This outbreak was driven by the 1E lineage of the South American I (SA-I) genotype that originated from the Amazon basin and spread from northern São Paulo into neighboring areas of western Minas Gerais and southern São Paulo, reaching areas without vaccine recommendations at the time (8). More recently, in March 2023, a *Callicebus nigrifrons* primate from Águas da Prata municipality (São Paulo state border with Minas Gerais) tested positive for YFV. Full genome analysis indicated a new introduction from the Brazilian mid-west region (9). Since September 2024, YFV cases were detected in humans and NTPs in São Paulo State in two distinct regions: Campinas surrounding area (Car) and Ribeirão Preto surrounding area (RPar). Here, we present phylogenetic findings from YFV genomes obtained in 2024-2025, indicating persistence of a new lineage that was likely introduced from the Midwest Brazil. Additionally, the growing number of cases in human and nonhuman primates confirms that the boundaries of YF circulation are expanding, giving rise to a new potential epidemic.

### The study

On September 18 and 24, 2024, liver samples of two NTP collected in Pedra Bela (Car) and Bragança Paulista (Car), respectively, tested positive for YFV by RT-qPCR (10) and/or immunohistochemistry. Liver samples were processed by Instituto Adolfo Lutz (IAL-SP), as previously described (4). In late December 2024, epizootic events (all *Alouatta caraya* primates) were confirmed in Ribeirão Preto (RPar), 200 Km away from the cases detected three months earlier. As of 13 February 2025, 21 NTP tested YFV RNA positive (Alouatta=12; 57.1%, Callithrix=5; 23.8%, Sapajus=3; 14.3%, 2 non informed) and YF activity has been detected in Pinhalzinho (Car), Socorro (Car), Campinas (Car), Colina (RPar), Ribeirão Preto (RPar), Serra Negra (Car) and Joanópolis (Car) municipalities, while 11 human cases were detected, with 8 death cases, all in Car (**Figure 1**). The story map of YFV dissemination can be visualized in <https://storymaps.arcgis.com/stories/968377dd8e86401ab4d3646a6f6a570a/preview>. For whole genome sequencing, cDNA synthesis, library preparation of six cases was performed at the Laboratório Estratégico from IAL-SP, using the Viral Surveillance Panel, Illumina. One additional human sequence from January 2025 generated using viral metagenomics (10) was also included. Consensus sequences were aligned using MAFFT. Maximum likelihood and Bayesian phylogenetic analyses were conducted using IQ-TREE2 and BEAST v.1.10.4, respectively (11,12). Sequences have been deposited in NCBI GenBank under accession numbers: PQ602526, PQ963937, PQ879115-PQ879118.

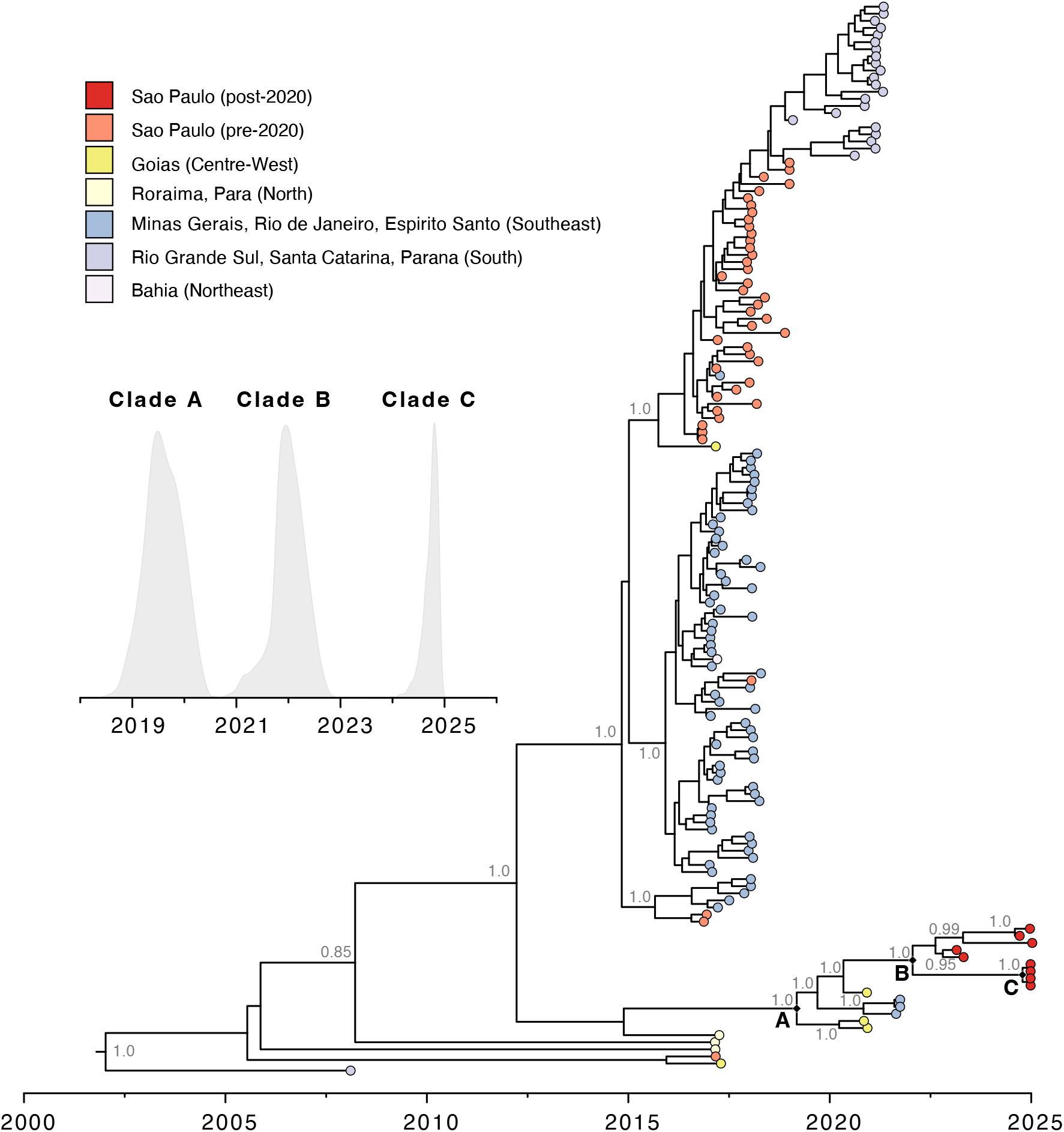

Phylogenetic analysis revealed that the 2024-2025 São Paulo sequences all clustered with maximum statistical support in a monophyletic cluster (Clade B) (**Figure 2**). This clade is most closely related to a sequence from Goiás (Centre-West Brazil), and genetically distinct from the viruses circulating during the 2016–2018 epidemic in São Paulo. The estimated date of the most recent common ancestor (TMRCA) of Clade B was around January 2022 (95% Bayesian credible interval, BCI, April 2021 to Aug 2022). We found that the recent 2024-2025 YFV genomes from RPar (Clade C), cluster together within Clade B. This suggests that following a reintroduction from Centre-West around early 2022, YFV has persisted and continued to spread within the São Paulo state. Our analyses also show that Clade B has split off from a more basal Clade A around August 2019 [95% BCI December 2018 to March 2020), which also contains additional sequences from Goiás and Minas Gerais states (Fig. 2).

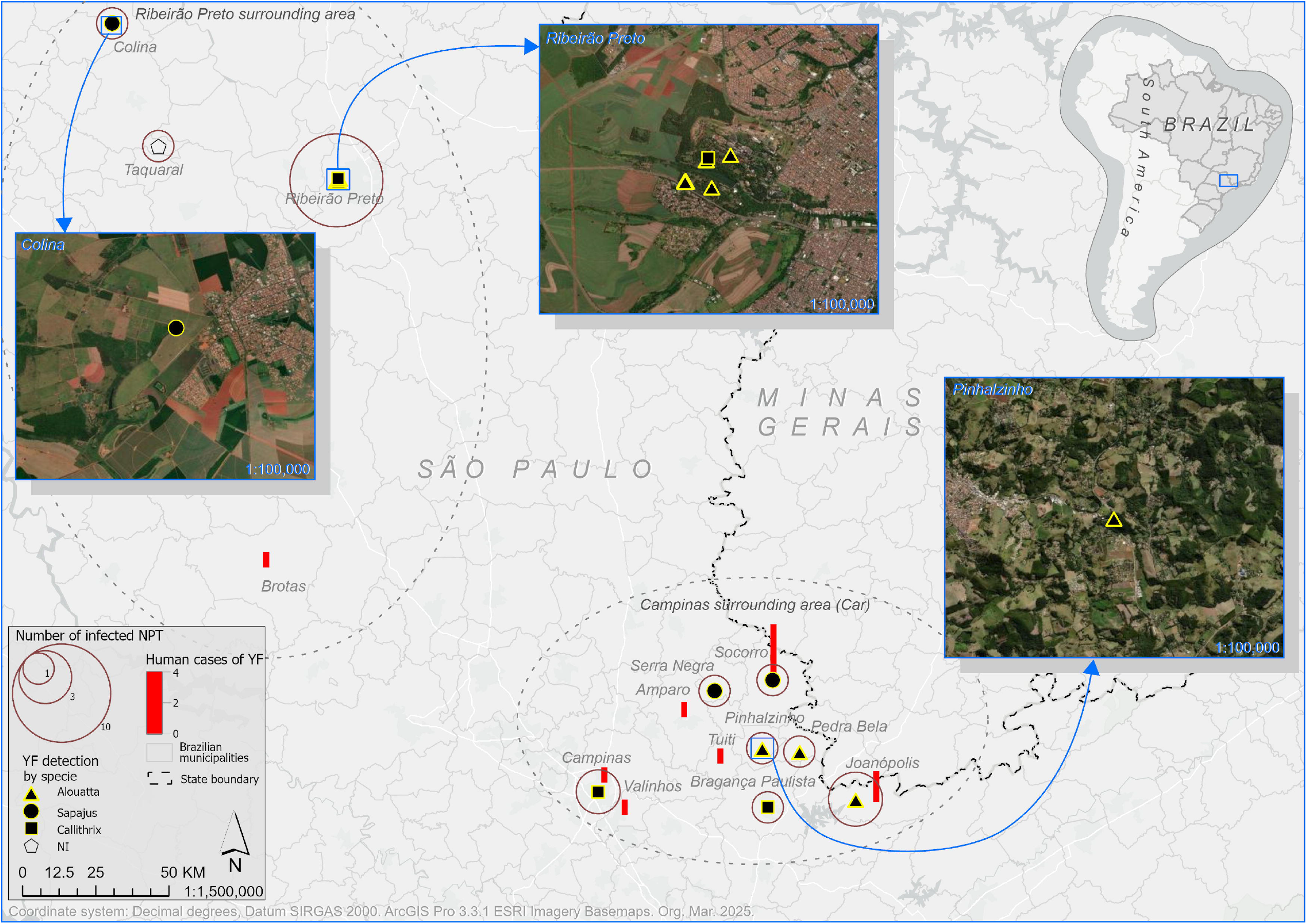

## Conclusion

Surveillance of sentinel NTPs remains the gold standard early warning system for detecting YFV circulation. However, it relies on the field and laboratory capacity of local authorities and prompt response of reference laboratories. Based on NTPs surveillance, vaccination strategies can be designed and targeted within and surrounding affected areas. In Brazil, YFV fatality rate in *Alouatta* genera is very high, and *Alouatta* populations have largely vanished during the 2016–2020 outbreak (5). Within the Car region, only half of the detected cases were in *Alouatta* genera, differently from the last epidemics. Instead, YFV was most frequently detected in *Sapajus* and *Callithrix*, two genera that are present in peri and urban sites which may therefore increase the risk of YFV reurbanization. Epizootic surveillance based on the early detection and notification of YFV in NTPs should be encouraged and strengthened across all regions.

## Data Availability

All data produced in the present work are contained in the manuscript

https://storymaps.arcgis.com/stories/968377dd8e86401ab4d3646a6f6a570a/preview

## Acknowledgments

We thank local veterinary authorities from the State of São Paulo for NTP samples collection, and Viviana Menezes for RNA extraction, the team from Center of Pathology, Adolfo Lutz Institute for sampling process, and the *Unidade de Vigilância de Zoonoses, Secretaria Municipal da Saúde Ribeirão Preto*. We also thank the Centro de Respostas Rápidas and the Laboratório Estratégico, Adolfo Lutz Institute, for NGS sequencing.

## Biographical Sketch

Dr. Cunha is a researcher at the Adolfo Lutz Institute. Her research interests include epidemiological surveillance and evolution of emerging and reemerging viruses with a focus on arboviruses and other zoonotic viruses.

## Funding

This study was supported by Fundo Especial de Saúde para Vacinação em Massa e Controle de Doenças (FESIMA), Secretaria de Estado de Saúde (SES) and CADDE project (**FAPESP 2018/14389-0)**

Authors declare no conflicts of interest

## Notes

### Competing Interest Statement

The authors have declared no competing interest.

